# Child exposure to animal feces and zoonotic pathogens in northwest Ecuador: A mixed-methods study

**DOI:** 10.1101/2025.08.28.25334671

**Authors:** Viviana Albán, April M. Ballard, Kelsey J. Jesser, Gwenyth O. Lee, Joseph N.S. Eisenberg, Daniel Garzón-Chávez, Gabriel Trueba, Bethany A. Caruso, Karen Levy

## Abstract

In low- and middle-income countries (LMICs), close cohabitation with animals and limited access to water, sanitation, and hygiene (WASH) infrastructure increase the risk of zoonotic enteric pathogen transmission to young children. This mixed-methods study combined (A) microbiological analysis of 120 animal fecal samples, and (B) go-along, semi-structured interviews with 35 mothers of children under two years across urban, intermediate, and rural communities in Ecuador to investigate: (Q1) What zoonotic enteric pathogens are present in animal feces and at what concentrations? (Q2) How are children exposed to animals and their feces? and (Q3) Which animals may serve as key sources of child? Microbiological analysis revealed high prevalence and concentrations of zoonotic pathogens, most commonly *E. coli* aEPEC (57%), *Salmonella* sp. (36%), and *E. coli* STEC (25%), with frequent co-infections (33%) and concentrations (4.97-9.29 log10 gc/g) often exceeding infectious dose thresholds. Qualitative findings showed risks from free-roaming animals, poor feces management, and frequent direct and indirect child–animal contact, often via caregivers and siblings. Triangulation identified chickens and dogs as major exposure sources due to their behaviors, proximity to children, and pathogen carriage. These findings highlight the need for targeted interventions to limit animal roaming, improve feces management, and increase caregiver awareness, while demonstrating the value of mixed-methods approaches to inform context-specific strategies for protecting child health in high-exposure environments.

## INTRODUCTION

In low- and middle-income countries (LMICs), animals serve as important sources of nutrition, integral components of agriculture, and significant economic assets in both rural and urban communities.[1, 2] However, cohabitation with animals and animal defecation in communal spaces, combined with inadequate access to water, sanitation, and hygiene (WASH) resources, can contribute to high concentrations of fecal contamination from animal sources in and around domestic environments.[3, 4]

Animal fecal contamination elevates the risk of zoonotic enteric pathogen transmission and poses a substantial threat to young children, whose developing immune systems render them particularly vulnerable to diarrhea and long-term impacts such as environmental enteric dysfunction, growth faltering, and cognitive deficits.[5–7] Vaccines and WASH interventions are common strategies to reduce enteric disease burdens. However, vaccines are not available to prevent diseases caused by all enteric pathogens,[8] and vaccination uptake can be low in LMICs for those that are available.[9] While WASH interventions can be effective to reducing risk of child diarrhea[10], large scale randomized trials have not resulted in the anticipated improvements in child health.[11–13] One reason may be because the interventions did not adequately address animal fecal contamination.[14, 15]

Animal fecal contamination of the environment in LMICs may be more extensive than human fecal contamination. Approximately 29.7 x 10^9^ kg of animal feces are produced globally every year, almost four times the amount of human feces.[16] Enteric pathogens like *Campylobacter* and *Salmonella*, Shiga toxin-producing *E. coli* (STEC), atypical Enteropathogenic *E. coli* (aEPEC), and *Cryptosporidium* parasites are known to be transmitted between humans and animals and are major contributors to the burden of human diarrheal disease.[1] Major transmission routes for enteric pathogens from animal feces include contamination of water through runoff, soils and crops via defecation or fertilizer use, and animal-derived foods. Additional pathways are unsafe feces disposal, contamination of household surfaces, and direct contact with fecal matter.[2]

Accurately characterizing human exposure to animal feces requires understanding both pathogen biology and the behaviors that mediate exposure.[2, 17] Collecting data on behaviors alongside pathogen presence and load in animal feces helps contextualize transmission pathways and identify animal sources of child risk. Such evidence is essential for designing effective, locally responsive interventions.

This mixed-methods study analyzes zoonotic enteric pathogen data and child and caregiver behaviors to characterize zoonotic enteric pathogen exposure pathways in children under two years of age along an urban-rural gradient in northwestern Ecuador. Our work addressed three research questions: (Q1) What zoonotic enteric pathogens are present in animal feces and at what concentrations? (Q2) How are children exposed to animals and their feces? (Q3) and Which animals may serve as key sources of child exposure to zoonotic pathogens, based on observed behaviors and fecal contamination? We also explored how these risks differ across the urban–rural gradient.

## METHODS

### Study design and setting

We used a convergent mixed methods design consisting of three components: (A) microbiological analyses of animal feces collected in and around participant households, to identify what zoonotic enteric pathogens are present and at what concentrations (Q1); (B) go-along semi-structured in-depth interviews (IDIs), a hybrid between participant observation and interviewing,[18, 19] to understand how children are exposed to animals and their feces (Q2); and (C) integration of microbiological and qualitative data to identify key animal sources of child exposure (Q3) (**Figure 1**). This approach combines microbiological evidence with contextual insights on child–animal and child–environment interactions to provide a holistic view of exposure pathways.

**Figure 1.**
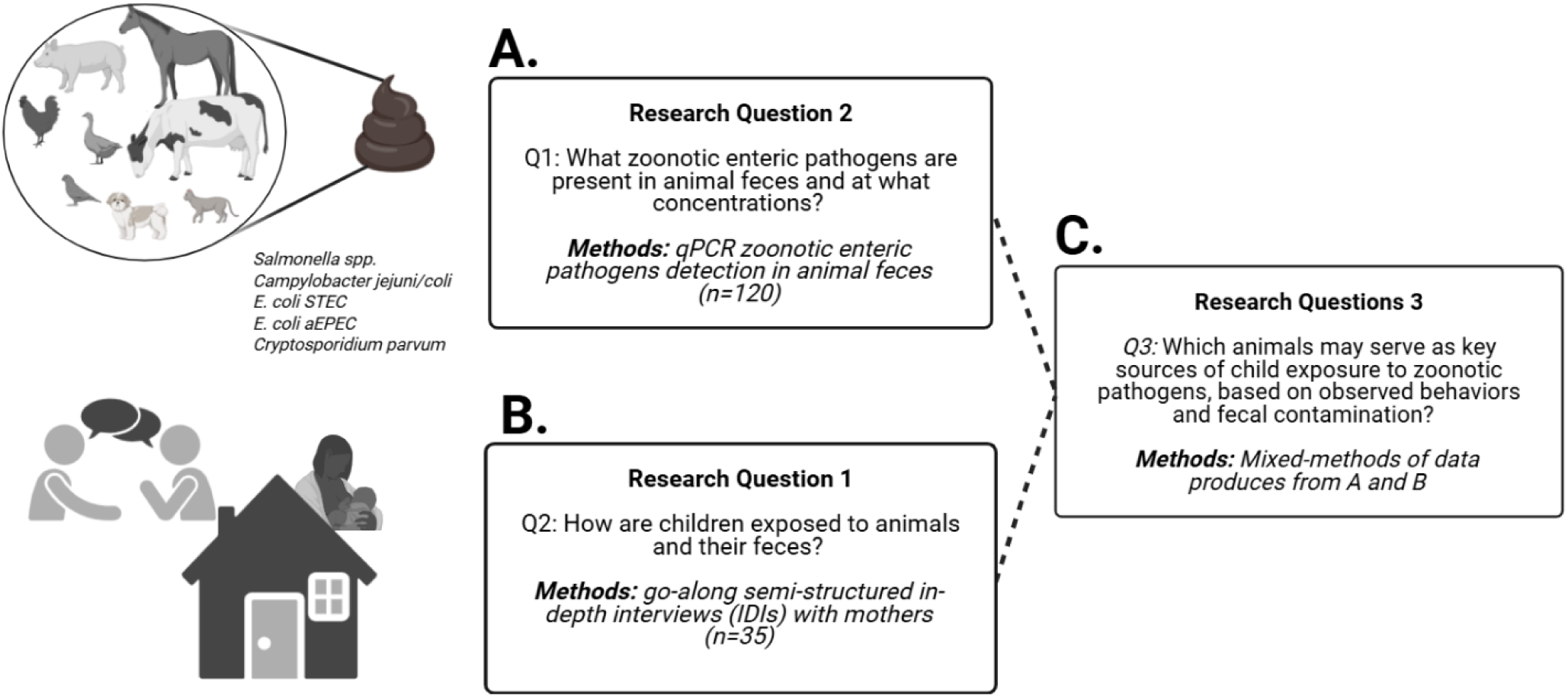
Integration of qualitative and microbiological methods (convergent mixed-methods design) to investigate zoonotic pathogen exposure in children. We used: **A.** Microbiological methods (qPCR) for the detection of zoonotic enteric pathogens to answer Q1 **B.** Qualitative methods, go-along semi-structured in-depth interviews (IDIs), to answer Q2. **C.** Triangulation of qualitative and microbial data to answer Q3.

**Figure 2:**
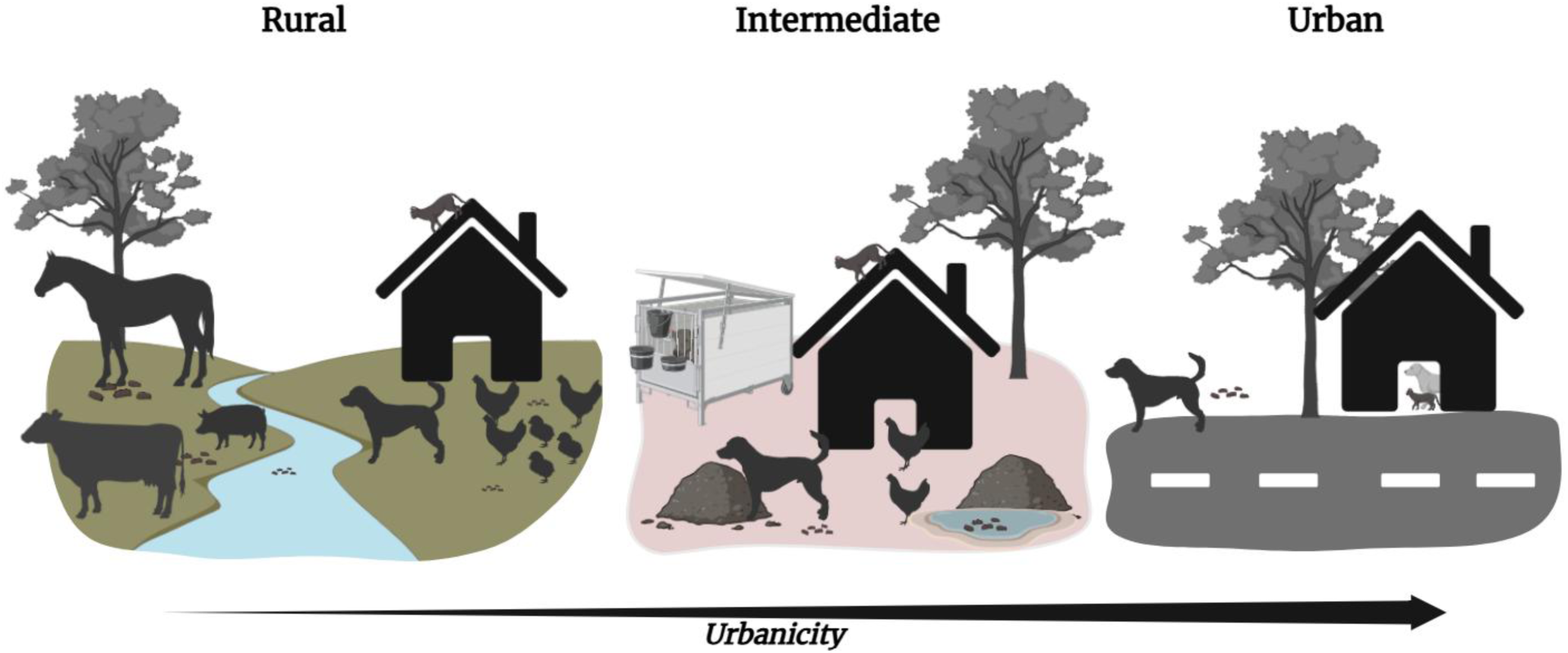
Conceptual illustration of animal fecal contamination across the urban-rural gradient. Illustration of changes in animal presence and the potential for environmental fecal contamination across the study areas. In rural areas (left), large animals are kept on farms across the river, while small animals roam freely around households. Animal feces from multiple animal types are commonly observed in the rural household environment. In the intermediate setting (center), both small and large animals are present near homes, though large animals are often kept in containment structures (e.g., animal pens). Animal feces from multiple animal types are commonly observed in the immediate household environment. In urban areas (right), fewer animals are present, and small animals, particularly pets, are more commonly kept in household patios or balconies. Animal feces, predominantly from dogs, are observed in the urban household environment.

We conducted this study between June and August 2019 Esmeraldas Province, northwestern Ecuador, across communities along an urban–rural gradient: (i) Esmeraldas city (‘***Urban’ community;*** population ∼150,000); (ii) Borbón (‘***Intermediate’ community;*** population ∼4,500), (iii) Maldonado (‘***Rural road’ community***; population ∼2,000), and (iv) Santo Domingo and Colón (‘***Rural river’ communities***; Santo Domingo: population ∼500; Colón: population ∼920). Santo Domingo and Colón are approximately 3.5 hours by boat from Borbón and are generally inaccessible by road, except for limited seasonal access. Due to small sample sizes, we combined the ‘Rural road’ and ‘Rural river’ communities into a single ***‘Rural’ communities*** category.

### Ethics

The Institutional Review Boards at Emory University (Atlanta, USA; STUDY00010353) and Universidad San Francisco de Quito (Quito, Ecuador; 2018-022M) approved the study protocol. Before data collection, we informed participants of the study aims, obtained written consent, and recorded interviews with their permission.

### Sample selection and size

To understand how children are exposed to animals and their feces, we recruited mothers of children under two who owned at least one animal. We focused on this age group because infants become increasingly mobile and vulnerable to environmental exposures,[7, 17, 20] and early childhood is a critical developmental period when repeated enteric infections can have lasting impacts on health and development.[5, 6]

Research assistants identified households with children under two and animal ownership by walking through communities. Using purposive sampling, we included households with diverse types and numbers of animals to represent each community. We aimed to conduct 30 IDIs (10 per community), with sample size informed by evidence that data saturation is typically reached within 9–17 qualitative interviews.[21]

### Data Collection and management Microbiological data

To identify what zoonotic enteric pathogens are present in what types of animal feces and at what concentrations, we collected 120 animal fecal samples from in and around the households of the mothers who were interviewed. Fecal samples for the following domestic animals were collected: chickens (n=28), dogs (n=21), cats (n=6), pigs (n=21), cows (n=14), horses (n=13), and other birds (ducks and parrots) (n=14).

For each animal fecal sample, we collected 5–10 g in sterile containers, stored them on ice, and transported them to nearby field labs. Within six hours, we aliquoted ∼1 g into four 1 ml cryovials and flash-froze them in liquid nitrogen. Samples were then transported to the Universidad San Francisco de Quito (USFQ) and stored at –80 °C.

We extracted genomic DNA from animal fecal samples using the PowerSoil® DNA Isolation Kit (MoBio Laboratories, Inc., Carlsbad, CA, US) following the manufacturer’s protocol. Quantitative PCR (qPCR) was used to detect and quantify aEPEC, STEC, *Salmonella* sp., and *Campylobacter* sp., while *Cryptosporidium* sp. was identified using an ELISA assay (**Appendix 1**). These pathogens were selected because they are commonly found in animal feces and represent important zoonotic risks.[1]

### Qualitative data

After obtaining consent, three trained Ecuadorian interviewers (two women, including author VAM, and one man) and a note-taker (non-Ecuadorian author AMB) conducted go-along, semi-structured IDIs. The guide covered animal ownership, environmental characteristics, child behaviors and interactions with animals, animal feces, and feces-contaminated soil, with probes used as needed. Mothers showed interviewers where animals lived and spent time; if observation was not possible, a traditional in-depth interview was conducted instead. Participants also completed a 10-question survey on maternal and child demographics, household water and sanitation, and animal ownership. With permission, interviews were audio recorded and lasted 15–60 minutes. Interviewers collected detailed notes, observations, and animal photographs (without human faces). After each interview, AMB and the team debriefed to review flow, discuss emerging themes, and document key observations to refine data collection in real time.[22]

### Data Analysis Microbiological data

Raw qPCR data were processed to determine presence/absence and abundance of targeted pathogens (Q1). Gene abundance was quantified relative to a mean standard curve, calculated from four curves run with each assay, and analyzed according to MIQE guidelines (**Appendix 1**).[23] *Cryptosporidium* sp. positivity was defined by an extinction rate cut-off (**Appendix 1**).

We performed all statistical analyses and data visualization using R version 4.4.1. Pathogen prevalence was calculated as the number of positive samples divided by the total tested. Chi-square tests assessed whether pathogen frequencies differed across communities and animal species, while Wilcoxon rank-sum tests compared pathogen concentrations (non-normally distributed) between communities. Finally, a probabilistic species co-occurrence analysis evaluated whether pathogen pairs occurred in fecal samples more or less often than expected by chance.[24]

### Qualitative data

To understand how children are exposed to animals and their feces (Q2), we analyzed observational and interview notes, audio transcripts, photographs, detailed interview summaries, and community profiles. We developed community profiles by compiling and synthetizing information from their sources of each community, including animal ownership, animal husbandry and animal health, feces exposure dynamics, child behaviors, and maternal perceptions and norms. We repeatedly reviewed the profiles and supporting data (audio, notes, photographs), wrote memos on emerging themes, and refined these themes through iterative re-reading and conceptualization. To enhance validity, authors AMB and VAM reviewed and discussed the thematic interpretation to ensure consistency and confirmability.

### Triangulation of qualitative and microbiological data

To identify which animals may serve as sources of child exposure to zoonotic pathogens (Q3), we triangulated findings from the microbiological analyses (Q1) and qualitative interviews (Q2). Specifically, we compared reported and observed patterns of child–animal interactions and environmental conditions with the prevalence of zoonotic pathogens detected in animal feces. This included linking behavioral themes, such as proximity of animals to living areas and child caregiving practices, to the detection of pathogens in different animal species across communities.

We integrated these streams of data to identify alignments and discrepancies between child behaviors and microbial contamination patterns, allowing us to draw connections between specific animals, exposure contexts, and settings. For example, animals frequently observed near children or in shared household spaces were examined for their pathogen carriage to assess potential child exposure.

Authors AMB and VAM jointly produced the thematic findings and iteratively reviewed and discussed them to ensure interpretive rigor and consistency.

To explore if and how animal-related risks vary along the urban-rural gradient for the three research questions, we stratified our results by community and identified overlap and divergence. For microbiological data, this included chi-square tests, Wilcoxon rank-sum tests, along with prevalence plots, boxplots, and summary tables. For qualitative data, this meant comparing key themes by iteratively reviewing notes, transcripts of audio recordings, photographs, summaries, and profiles.

## RESULTS

### Sample characteristics

The final qualitative sample included 35 mother–child dyads. Most households owned dogs (63%) and/or cats (60%) (Table 1). Animal ownership diversity increased with rurality: urban households owned 1–2 species, intermediate 1–4, and rural 1–6. Mothers averaged 26 years of age (range: 18–43) and children 13 months (range: 2–23) (**Table S2**). Additional household and demographic details are provided in **Table S2**.

**Table 1:**
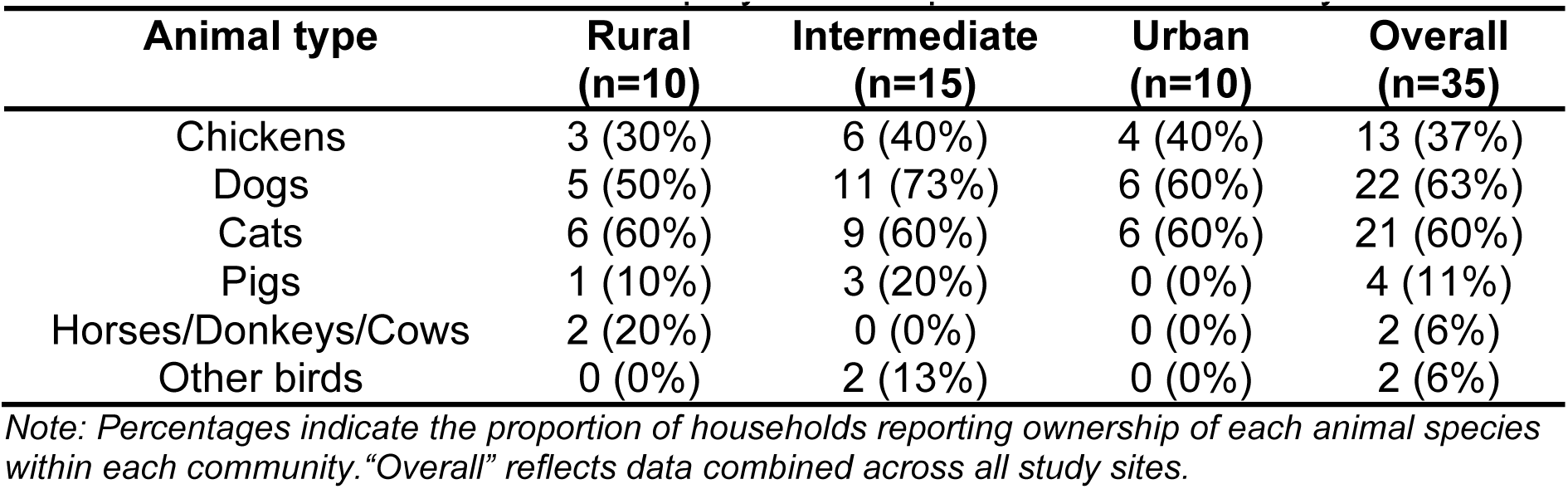
Household animal ownership by animal species and community.

| **Animal type** | **Rural**<br>**(n=10)** | **Intermediate**<br>**(n=15)** | **Urban**<br>**(n=10)** | **Overall**<br>**(n=35)** |
| --- | --- | --- | --- | --- |
| Chickens | 3 (30%) | 6 (40%) | 4 (40%) | 13 (37%) |
| Dogs | 5 (50%) | 11 (73%) | 6 (60%) | 22 (63%) |
| Cats | 6 (60%) | 9 (60%) | 6 (60%) | 21 (60%) |
| Pigs | 1 (10%) | 3 (20%) | 0 (0%) | 4 (11%) |
| Horses/Donkeys/Cows | 2 (20%) | 0 (0%) | 0 (0%) | 2 (6%) |
| Other birds | 0 (0%) | 2 (13%) | 0 (0%) | 2 (6%) |

*Note: Percentages indicate the proportion of households reporting ownership of each animal species within each community. "Overall" reflects data combined across all study sites.*

**Table 2:**
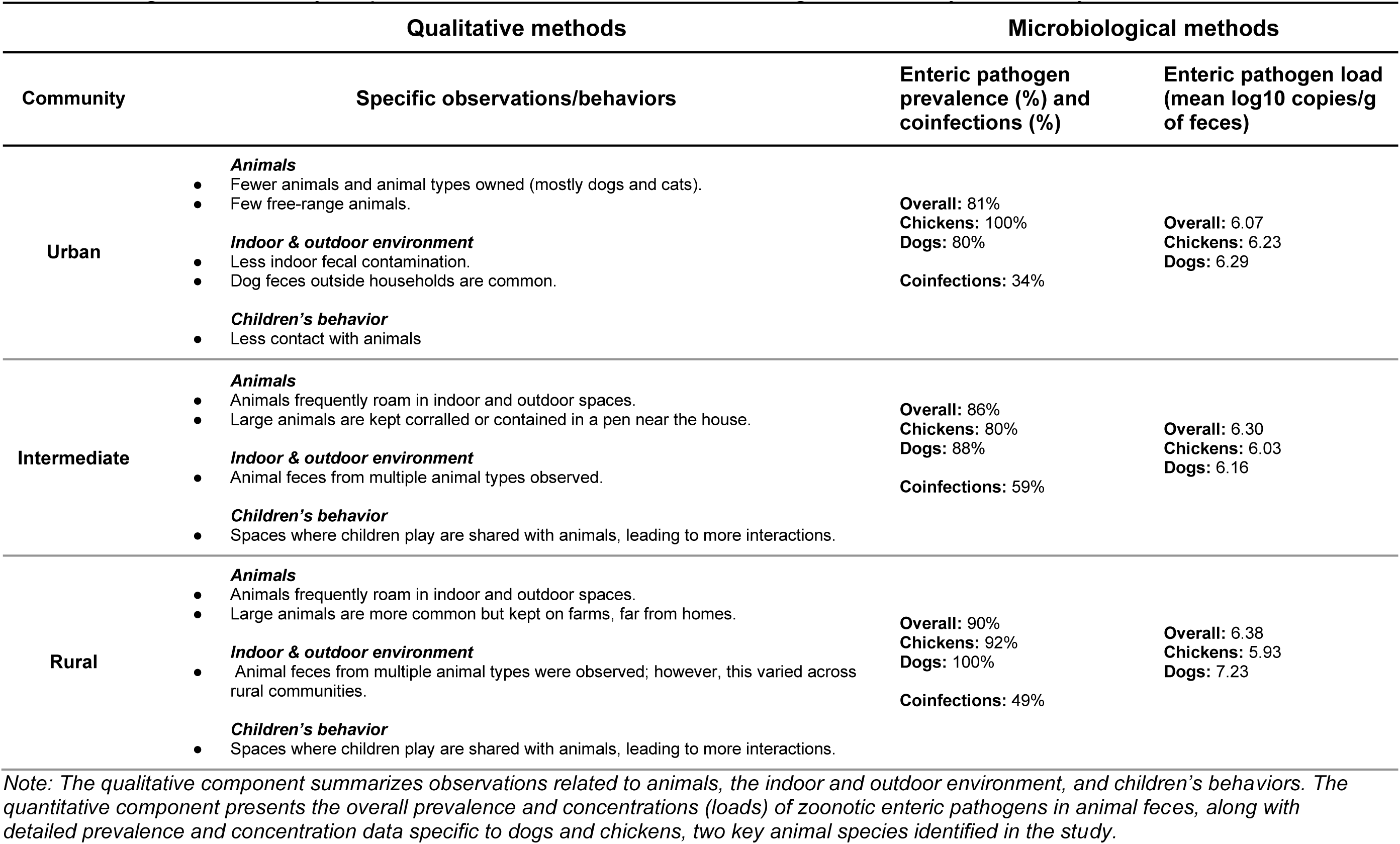
Integrated summary of qualitative observations and microbiological results by community.

### Q1: What zoonotic enteric pathogens are present in animal feces and at what concentrations?

#### Zoonotic enteric pathogens were frequently detected in animal feces and often were present at high concentrations

Overall, zoonotic pathogens were detected in 106 of 120 samples (88%). The proportion positive for at least one pathogen varied by animal species: chickens 89% (25/28), dogs 90%(19/21), cats 83% (5/6), pigs 86% (18/21), cows 93% (13/14), horses 85% (11/13), and other birds (ducks and parrots) 76% (13/17). The overall prevalence of enteric pathogens varied significantly by animal species (X^2^ (24, N = 120) = 81.12, p < 0.0001). Across all animals, the diarrheagenic *E. coli* pathotype aEPEC was the most common enteric pathogen, detected in 57% of samples and present across all animal species.

*Salmonella* sp. was detected in 36% animal feces, primarily in pigs (62%), cows (57%), and cats (50%). The diarrheagenic *E. coli* pathotype STEC was found in 25% of all animal feces, with carriage most frequently detected in cows (71%).

*Campylobacter coli/jejuni* was present in 19% of animal feces, predominantly in chickens (43%). *Cryptosporidium* sp. was least common, detected in 18% of animal feces, with horses being notable carriers (77%) (**Figure 3A**).

**Figure 3:**
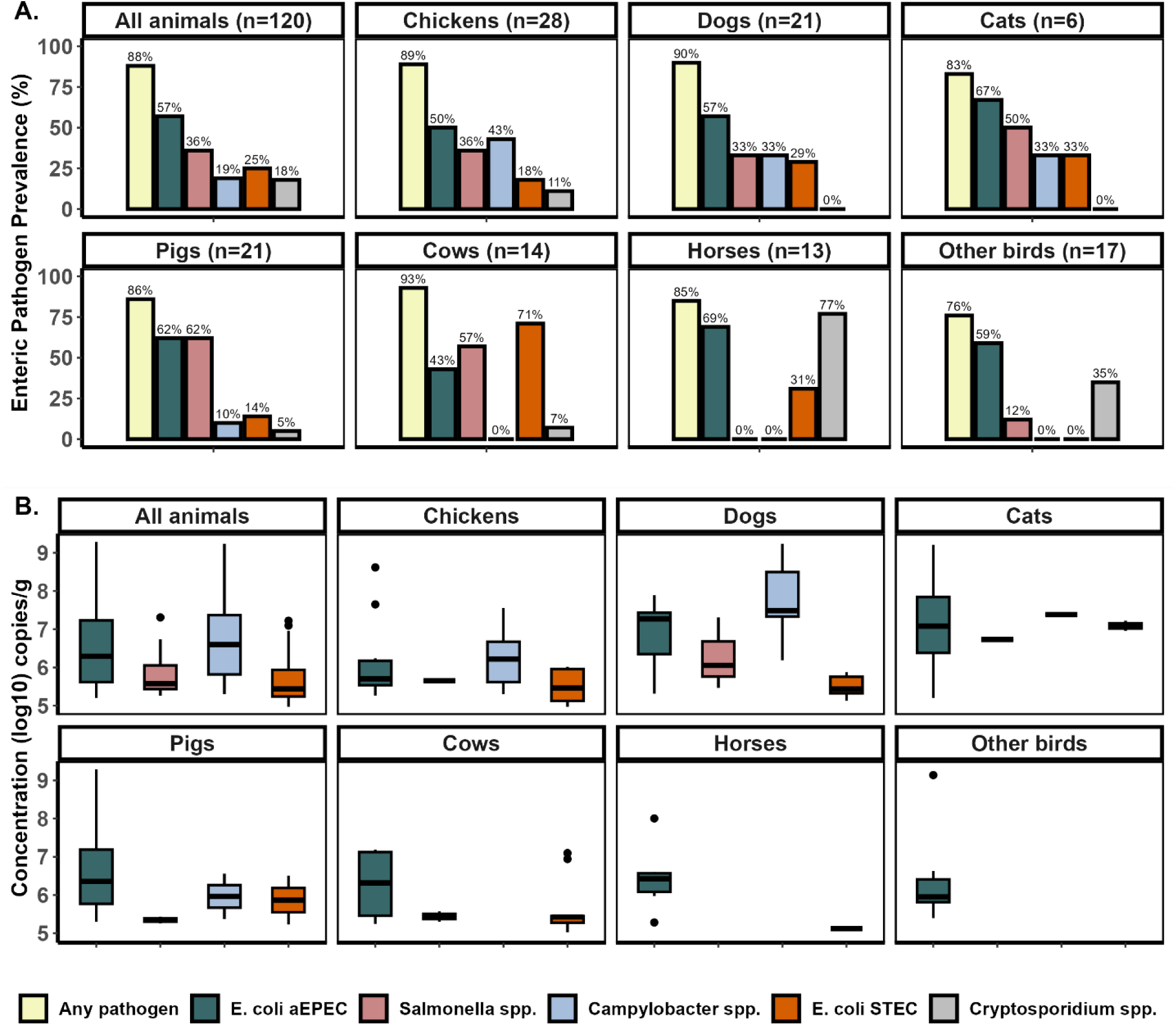
Prevalence and concentration of zoonotic enteric pathogens in animal feces by animal species. **A**. Bar plots show the prevalence (%) of each enteric pathogen detected in animal fecal samples, stratified by site type. Each bar represents the proportion of positive samples for a given pathogen. Pathogens include aEPEC, *Salmonella* sp., *Campylobacter* sp., STEC, and *Cryptosporidium* sp., with “Any pathogen” indicating detection of at least one pathogen in the sample. Bar height and color reflect pathogen-specific prevalence. **B.** Boxplots showing the concentration of pathogens (log10 copies/g) detected in animal fecal samples by animal species. Each box represents the interquartile range (IQR) with the median indicated by a horizontal line. *Cryptosporidium* sp. concentrations are not shown because the ELISA assay used only allowed for presence/absence detection.

Across positive samples, pathogen concentrations spanned 4.97 and 9.29 log10 gc/g of feces for all targets. Mean aEPEC concentrations were broadly similar across animal species (chickens 6.16, dogs 6.86, cats 7.14, pigs 6.72, cows 6.27, horses 6.45, and other birds: 6.34 log10 gc/g). Cats and dogs showed higher mean concentrations of *Salmonella* sp.(6.73 and 6.27 log10 gc/g) and *Campylobacter coli/jejuni* (7.75 and 7.38 log10 gc/g) than in other animal species (**Figure 3B**). STEC mean concentrations were also higher in cats (7.09 log10 gc/g) than in other animal species. However, pairwise Wilcoxon rank-sum tests indicated no significant differences in pathogen concentrations between animal species for any enteric pathogen (all p-values > 0.05). Coinfections, defined as the detection of more than one enteric pathogen in a single animal fecal sample, occurred in 48% of samples and were common across animal species; chickens (57%) and horses (62%) had the highest coinfection rates (**Figure 5A**). We identified 11 enteric pathogen co-infection patterns, the most common pattern being *Salmonella* sp. + STEC (21%), followed in order by aEPEC + *Salmonella* sp. (19%), aEPEC + *C. parvum* (16%), and aEPEC + *C. jejuni/coli* (14%) (**Table S3**). A probabilistic co-occurrence model indicated that most observed co-occurrences did not deviate from random expectation (**Figure S2**).

Detection of potentially zoonotic enteric pathogens was common in animal feces from all communities (**Figure 4A**), although a non-significant decreasing trend was observed with increasing urbanicity (Chi-square (8, N = 120) = 2.2151, *p* = 0.9737). aEPEC and *C. jejuni/coli* were found at concentrations of ∼1 log higher in the rural communities compared to the intermediate and urban communities. In contrast, *Salmonella* sp, and *E. coli* STEC had concentrations ∼1 log higher in the intermediate community than in rural and urban communities (**Figure 4B**). We found no significant differences in enteric pathogen concentrations across communities.

**Figure 4:**
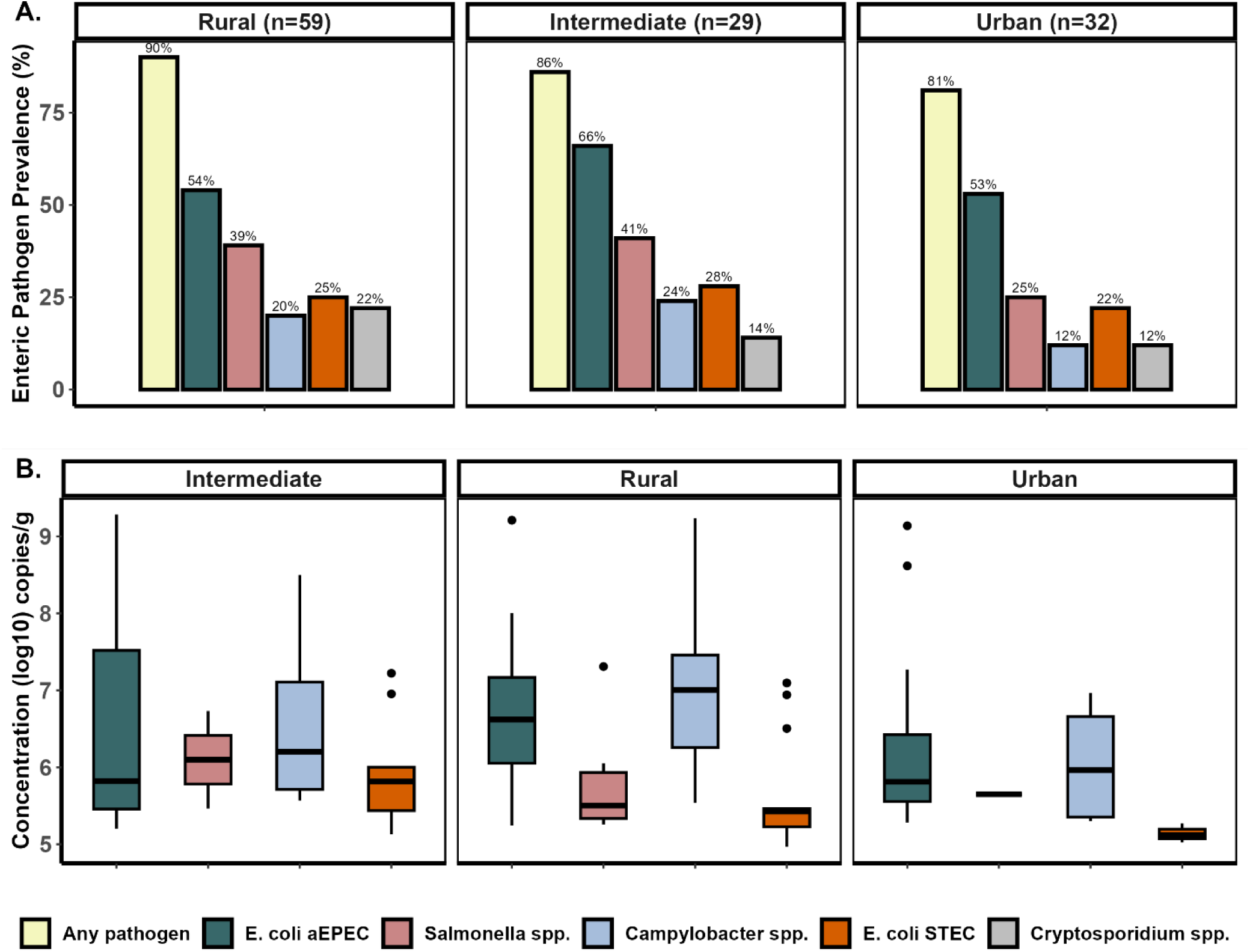
Abundance and coinfection patterns of zoonotic enteric pathogens in animal feces by community. **A.** Bar plots show the prevalence (%) of each enteric pathogen detected in animal fecal samples, stratified by community. Each bar represents the proportion of positive samples for a given pathogen. Pathogens include aEPEC, *Salmonella* sp., *Campylobacter* sp., STEC, and *Cryptosporidium* sp., with “Any pathogen” indicating detection of at least one pathogen in the sample. Bar height and color reflect pathogen-specific prevalence. **B.** Boxplots showing the concentration of pathogens (log10 copies/g) detected in animal fecal samples by community. Each box represents the interquartile range (IQR) with the median indicated by a horizontal line. *Cryptosporidium* sp. concentrations are not shown because the ELISA assay used only allowed for presence/absence detection.

Coinfection with multiple pathogens was most common in intermediate (59%) and rural (49%) communities, compared to the urban (34%) community; however, these differences were not statistically significant. (**Figure 5B**).

**Figure 5:**
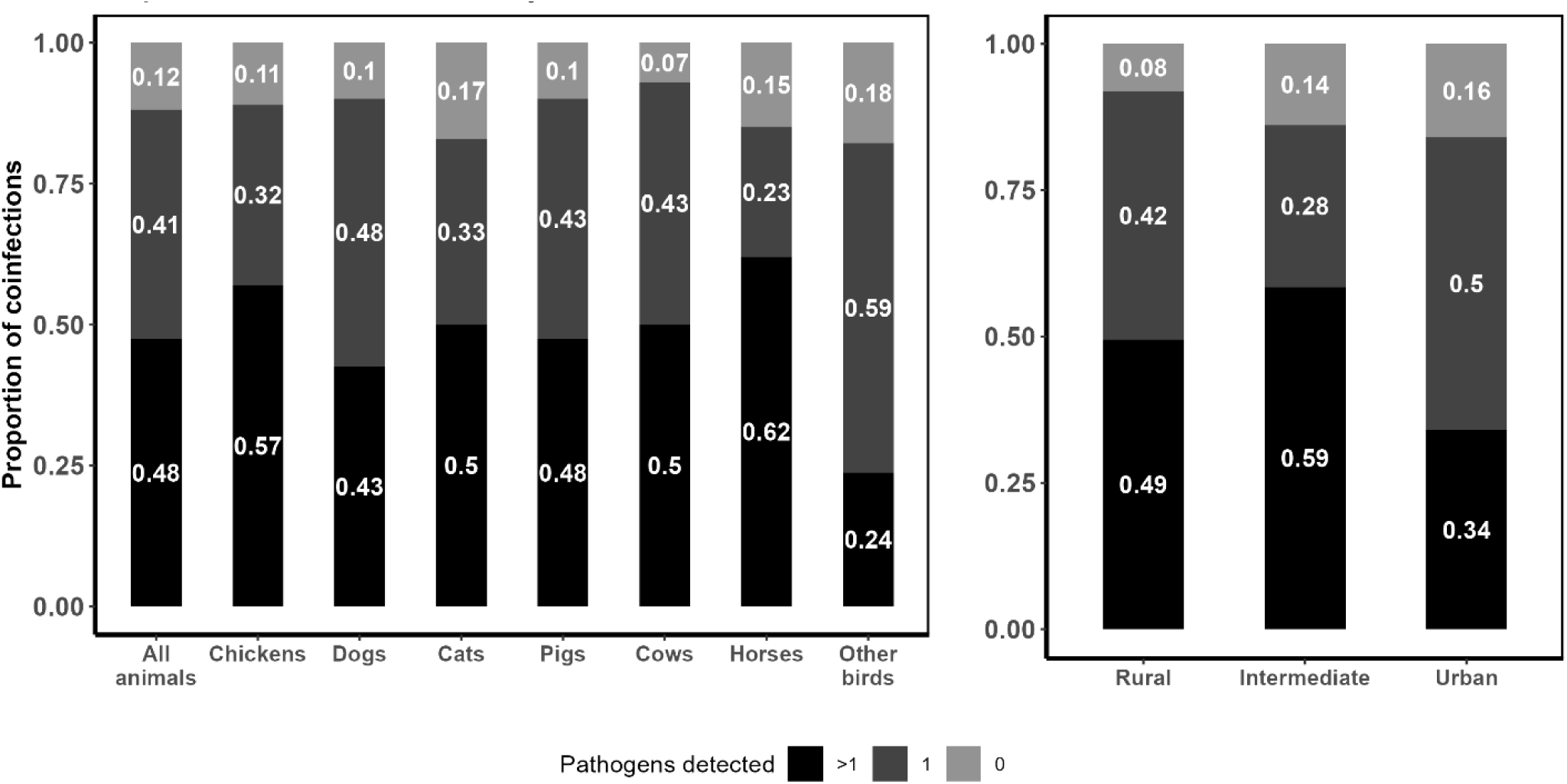
Proportion of coinfections, single infections or no infection with zoonotic enteric pathogens in animal fecal samples by animal species and community. Stacked bar plots depicting the proportion of coinfections (*black*), single infections (*dark gray*) and no infections (*light gray*) in animal fecal samples, stratified by animal species and community.

### Q2: How are children exposed to animals and their feces?

#### Widespread animal presence and free-roaming practices increased opportunities for child-animal contact

Widespread animal presence and free-roaming practices created frequent opportunities for child–animal contact. Dogs, cats, and free-range chickens commonly roamed within and between households, often sharing domestic spaces used for food preparation, laundry, and child play. In one intermediate community, for example, a mother allowed her dog, cat, and chickens to move freely through the backyard, house, and neighborhood; the same areas where animals ate, slept, and defecated were also used by children and caregivers. In contrast, larger animals such as pigs, cows, and horses were generally penned or kept away from houses.

Animal presence and roaming patterns varied by community. Urban households generally owned fewer animals and restricted their movement with fenced yards, closed doors, and by tying dogs near entrances as guards. In contrast, intermediate and rural households owned more animals and imposed fewer barriers, allowing freer movement. Urban mothers also cited safety concerns and local norms, such as keeping doors closed, as reasons for limiting animal entry.

#### Environmental fecal contamination was common outdoors and occasionally indoors, especially from dogs and chickens

Although mothers often downplayed indoor contamination, observations revealed instances in kitchens and living spaces, for example, parrot feces under a chair after apparent cleaning, and dog feces in a kitchen where puppies were kept. Cats were widespread but contributed little visible contamination due to their burying behavior.

Outdoor fecal contamination varied more across communities than indoor contamination, which was generally infrequent. Community practices and household routines appeared to shape these differences. In one rural community, larger animals were kept on farms away from homes, and social norms encouraged families to clean up feces promptly, as neighbors would notice if waste was left in shared spaces. In another rural community, where animals were kept closer to homes and no such norms were described, animal feces were more often observed near households. For example, in one household, a mother fed rice to free-range chickens outside her door each evening to corral them, contributing to feces accumulation near the entrance.

#### Child–animal and family interactions created frequent exposure opportunities

Children under two often had direct contact with dogs and cats, and sometimes chickens, while playing or crawling in shared indoor and outdoor spaces. Hand-to-mouth and object-to-mouth behaviors were common; in one rural household, a child was observed crawling where chickens had just defecated, chasing them, and putting objects, and even feces, into their mouth. Such direct contact was less common among urban children, reflecting fewer animals and stricter management practices.

Beyond direct contact, older siblings and caregivers were important indirect exposure pathways, particularly in rural settings. Children frequently had close contact with family members who handled animals or contaminated surfaces, for example, a sibling played with chickens and dogs and then touched the face and mouth of a child under two. Fathers, who often worked with farm animals, also represented potential sources of transmission. In urban households, less frequent animal contact among family members reduces these indirect risks.

### *Q3:* Which animals may serve as key sources of child exposure to zoonotic pathogens, based on observed behaviors and fecal contamination?

Chickens and dogs emerged as key sources of child exposure to zoonotic pathogens, supported by both qualitative observations and microbiological analyses. These animals were major contributors to persistent fecal contamination in household environments, as mothers reported, and observations confirmed, frequent child interactions with them. Free-ranging practices further led to widespread dog and chicken feces around households, heightening exposure risks.

Microbiological testing confirmed chickens and dogs as primary exposure sources: 89% of chickens (25/28) and 90% of dogs (19/21) carried at least one enteric pathogen. aEPEC, *Salmonella* sp., and *Campylobacter* sp. were each detected in >30% of samples (Figure 3B), with mean concentrations in dogs reaching 6.1 log10 gc/g for *Salmonella* sp. and 7.5 for *Campylobacter* sp. (**Figure 3**). Co-infections were frequent, affecting 57% of chickens and 43% of dogs, most commonly aEPEC with *Campylobacter* sp. and *Salmonella* sp. with STEC (**Table S3**).

## DISCUSSION

This mixed-methods study examines how animal fecal contamination, caregiver and child behaviors, and animal management practices shape young children’s exposure to zoonotic enteric pathogens along an urban–rural gradient in northwestern Ecuador. By integrating microbiological data with qualitative observations and maternal reports, we addressed three questions: (Q1) which pathogens are present and at what concentrations, (Q2) how children are exposed to animals and their feces, and (Q3) which animals serve as key exposure sources.

Our findings highlight persistent and widespread exposure pathways that reflect the interplay of community norms, household practices, and child-environment interactions.

### High diversity and high concentrations of zoonotic enteric pathogens highlight a persistent risk (Q1)

The diversity of zoonotic enteric pathogens detected across animal species, along with high coinfection rates, highlights children’s risk of simultaneous exposure to multiple pathogens. Some host–pathogen patterns aligned with expectations, such as widespread detection of aEPEC across species,[25] and STEC in cattle, consistent with their role as a primary reservoir.[26–29] Other patterns were less expected. For example, in our sample, *Salmonella* sp. was rarely found in chicken feces, despite poultry being a widely recognized reservoir.[30, 31] This may reflect the predominance of backyard, low-density, free-ranging poultry systems in our sample, which may reduce transmission compared to intensive operations.[32] However, more concentrated poultry production does occur in the region,[33, 34] and pathogen dynamics in those settings may differ from those observed here.

Co-infections involving multiple enteric pathogens were common in animal feces, especially combinations of *Salmonella* sp. with STEC and aEPEC. Such coinfections may exacerbate child health risks by increasing environmental pathogen loads and prolonging shedding.[35] Consistent with this, TaqMan Array Card testing of infant feces at six months showed that 72% (n=276) of children carried more than two pathogens,[36] reinforcing the likelihood of cumulative exposure in early childhood. Similar findings from rural Bangladesh link multiple infections in child feces to greater stunting risk.[37]

Pathogen concentrations in animal feces were consistently high and often exceeded estimated human infectious doses. *Salmonella* sp. reached up to 7 log10 gc/g, *Campylobacter* sp. 6.69 log10 gc/g, aEPEC 6.53 log10 gc/g, and STEC 5.69 log10 gc/g, while infectious doses range from 2.8 to 10⁴ cells for *Salmonella* sp.,[38] fewer than 10³ cells for *Campylobacter* sp.,[39] and as few as 50 cells for E. coli STEC.[40] These high concentrations suggest that even brief or indirect contact with animal feces or indirect contact may be sufficient to initiate infection, particularly in young children with developing immune systems and limited hygiene resources. At the same time, we note that risk depends on the degree to which fecal material is attenuated before contact (natural die-off, dilution/dispersion, or handling), which we did not quantify here.

Although the urban community in our study region generally had better WASH infrastructure and lower livestock densities, these advantages did not significantly reduce the presence of animal feces in the environment. Pathogen prevalence and concentrations in animal feces were only modestly lower in the urban community, and the differences were not statistically significant, reinforcing that exposure is shaped less by geography and more by the interplay of community, household, and child-level factors, including behaviors at all these levels. We also found that pathogen distribution varied significantly across animal species, suggesting that the type of animal present or interacted with may be more critical for exposure risk than location along the urban–rural spectrum.

### Daily household and caregiving behaviors sustain multiple exposure pathways (Q2)

Animal management practices and caregiver behaviors across communities created sustained opportunities for zoonotic pathogen exposure. Chickens and dogs were commonly present in both outdoor and indoor spaces, including food preparation, laundry, and child play areas, critical sites where fecal contamination directly threatens child health. Prior research in Bangladesh[4] and Ethiopia[41] supports these concerns, showing associations between animal feces in domestic settings and adverse health outcomes such as environmental enteropathy, stunting, and impaired growth.

Children frequently engaged in high-risk behaviors, including crawling and playing on contaminated surfaces, frequent hand-to-mouth contact, and geophagy. In northwestern Ecuador,[42] children averaged 2.5 hand-to-mouth contacts and 0.38 oral contacts with soil or garbage per hour, with similar behaviors documented in Bangladesh,[43] Zimbabwe,[7] and Perú.[44] These patterns emphasize the near-constant nature of environmental contact in early childhood and its role in sustaining high exposure to zoonotic pathogens. [2, 14, 45, 46]

Interactions between children and other household members also emerged as an important, yet overlooked, transmission pathway.[17, 47] Although many mothers reported keeping infants in animal-free areas, we observed frequent physical contact with caregivers, siblings, and others, creating opportunities for indirect fecal–oral transmission through contaminated hands. Evidence from Ethiopia[48] and Bangladesh[49] shows *E. coli* and ruminant fecal markers on mothers’ and children’s hands, with strong mother–child correlations, and mothers often reported manually removing animal feces. Our prior work in northern Ecuador[36] similarly found widespread *E. coli* contamination on maternal and child hands, reinforcing the significance of interpersonal pathways in sustaining exposure.

### 4.3 Dogs and chickens are priority animal species for intervention (Q3)

Among all animal species, dogs and chickens emerged as key animal sources of child exposure to zoonotic pathogens. Chicken feces was particularly ubiquitous and not often removed, likely due to their small size, relatively low odor, and tendency to be overlooked by household members.[50] These findings mirror patterns in Zambia[51] and Bangladesh[4] where household poultry ownership is common but feces management is not. In contrast, feces from larger animals (e.g., pigs, cows, horses) were less commonly found near households, largely due to practices that physically restrict their movement (e.g., penning or tethering). This distinction between free roaming versus contained animals appears to be a key determinant of environmental contamination patterns and subsequent exposure risk for children, as small animal feces might create numerous and repeated opportunities for fecal–oral transmission. These results contribute to the scientific literature calling for the identification of hot-spots of human contact with animals,[2] making dogs and chickens particularly important targets for targeted interventions in this study area.

A key strength of this study is its convergent mixed-methods design, which integrated in-depth behavioral observation with microbiological testing to provide a more complete picture of everyday exposure pathways. Sampling communities along an urban–rural gradient further supports the relevance of findings for intervention design across diverse settings. Limitations include modest sample sizes and cross-sectional sampling, which limit generalizability and preclude assessment of temporal variation. In addition, we did not account for attenuation of fecal material before contact (e.g., die-off, dilution, removal) or pathogen viability, so qPCR gene copies may overestimate infectious load.

Children’s exposure to zoonotic enteric pathogens in animal feces is shaped by interconnected animal, environmental, microbiological, and behavioral factors. Key risks include the presence of animals in and around households, frequent child– animal contact, and high pathogen burdens in animal feces. Although rural–urban context influences these dynamics, geography alone does not determine risk.

Improving child health and reducing stunting therefore requires strategies that go beyond traditional WASH, including restricting animal roaming, promoting regular feces removal with clear community guidance, creating safe animal-free play spaces, and reinforcing caregiver and sibling hygiene practices to limit indirect transmission.

## ACKNOWLEDGEMENTS

We acknowledge the families who enrolled in the study and the local field staff who conducted the interviews and surveys and supported the transportation of samples.

## DATA AVAILABILITY STATEMENT

The datasets and R code supporting this study are available on GitHub at: https://github.com/valban337/Mixed-methods-animal-exposure-study,

