## Supplementary Information for "Child exposure to animal feces and zoonotic pathogens in northwest Ecuador: A mixed-methods study"

**Appendix 1: qPCR and ELISA methods.**

**qPCR assays.** Assay gene targets along with primers, probes, and standard gblock (synthetic gene fragments containing all enteric pathogens target genes) sequences are presented in **Table S1**. We generated standard curves from serial 10-fold dilutions of a gBlock standard containing the target DNA sequences, with concentrations ranging from $10^6$ to $10^2$ gene copies per reaction (**Figure S1**). We included a standard curve on each qPCR assay plate and prepared a fresh gblock dilution series daily.

We ran standards and samples duplicate on a real-time PCR system (CFX96, Bio-Rad, USA) in a final reaction volume of 20 uL. Each reaction included 10 ul 2X Taqman® Universal PCR Master Mix (Applied biosystems, Life Technologies Corporation, Carlsband, CA, US), 1 uM of each forward and reverse primer, 0.1 uM of probe, and 4 ul of DNA template. Cycling conditions were as follows: 50°C for 2 min, 95°C for 10 min, and 40 cycles of 95°C for 15 sec, and 55°C for 1 min (for *invA*[20], *eae*[21], *stx1*[21] and *stx2*[21]) / 60°C for 1 min (for *bfpA*[21] and *cadF*[21]). We included non-template controls (NTCs) and negative extraction controls (NECs) in each run. We assessed qPCR inhibition by spiking each DNA extract with1 x $10^6$ copies of a gblock containing a 220-bp artificial inhibition control (IC) sequence.[22] We amplified the IC using a SYBR green qPCR assay and confirmed specificity by melt curve analysis. No inhibition of qPCR amplification was detected.

We defined the assay limits of detection (LoD) as the lowest standard concentration that met two criteria: a standard deviation <1 for replicates and >95% replicate detection. We calculated assay limits of quantification (LoQ) using the LoD Cq value and its standard deviation (σ) as follows: CtLoQ = CtLoD – 2(σLoD). We summarized assay LoD, efficiency, linear dynamic range, and standard curve $R^2$ values and slopes for each assay in **Table S1**. NTCs and NECs included on each plate showed no amplification at 40 cycles.

We considered a sample Detectable and Quantifiable (DQ) if the target amplified in both duplicate qPCR reactions and the mean Cq value was >LoQ and between the highest and lowest dilutions on the standard curve. We classified samples Detectable but Not Quantifiable (DNQ) or Not Detected (ND) otherwise.[26] DQ and DNQ results were considered positive (present), while ND results were considered negative (absent) for prevalence analyses.

**ELISA assays.** We determined the presence or absence of *Cryptosporidium* spp. using the RIDASCREEN® *Cryptosporidium* enzyme immunoassay, following the manufacturer's instructions. We included positive and negative controls provided in the kit on each ELISA plate. We established a cut-off value by adding 0.15 extinction units to the negative control measurement. A sample was considered positive for *Cryptosporidium* if its extinction rate exceeded the cut-off by more than 10%.

### Supplementary Tables

**Table S1.** Primers and probe sequences for enteric pathogen gene targets and analytical performance of qPCR assays.

| Enteric pathogen | Gene | Primers | Efficiency (%)[1] | R2[1] | Slope | Y-intercept | LOD[2] *(equiv. no. of copies)* | Reproducibility[3] *(Low- and high-concn. CV (%))* |
|---|---|---|---|---|---|---|---|---|
| *Salmonella* | *invA*[12] | **F:** GCTGCTTTCTCTACTTAAC<br>**R:** GTAATGGAATGACGAACAT<br>**P:** FAM-CATCACCATTAGTACCAGAATCAGT-BHQ1 | 94.57 | 0.99 | -3.46 | 46.42 | $10^5$ (100) | 0.99-1.42 |
| *C. jejuni/coli* | *cadF*[13] | **F:** CTGCTAAACCATAGAAATAAAATTTCTCAC<br>**R:** CTTTGAAGGTAATTTAGATATGGATAATCG<br>**P:** FAM-CATTTTGACGATTTTTGGCTTGA-BHQ1 | 93.46 | 0.99 | -3.49 | 48.02 | $10^5$ (100) | 0.54-1.05 |
| *E. coli* (aEPEC) | *eae*[13] | **F:** CATTGATCAGGATTTTTCTGGTGATA<br>**R:** CTCATGCGGAAATAGCCGTTA<br>**P:** FAM-ATACTGGCGAGACTATTTCAA-BHQ1 | 95.02 | 0.99 | -3.45 | 47.26 | $10^5$ (100) | 0.72-2.90 |
| *E. coli* (aEPEC) | *bfpA*[13] | **F:** TGGTGCTTGCGCTTGCT<br>**R:** CGTTGCGCTCATTACTTCTG<br>**P:** FAM-CAGTCTGCGTCTGATTCCAA-BHQ1 | 96.72 | 0.99 | -3.40 | 44.71 | $10^5$ (100) | 0.39-2.93 |
| *E. coli* (STEC) | *sxt1*[13] | **F:** ACTTCTCGACTGCAAAGACGTATG<br>**R:** ACAAATTATCCCCTGWGCCACTATC<br>**P:** FAM-CTCTGCAATAGGTACTCCA-BHQ1 | 93.3 | 1.00 | -3.50 | 48.00 | $10^5$ (100) | 0.49-1,00 |
| *E. coli* (STEC) | *stx2*[13] | **F:** CCACATCGGTGTCTGTTATTAACC<br>**R:** GGTCAAAACGCGCCTGATAG<br>**P:** FAM-TTGCTGTGGATATACGAGG-BHQ1 | 92.24 | 1.00 | -3.52 | 46.21 | $10^5$ (100) | 0.22-1.26 |
| | IC[14] | **F:** CTAACCTTCGTGATGAGCAATCG<br>**R:** GATCAGCTACGTGAGGTCCTAC | | | | | | |

*All analysis was based on four standard curves per target

1 The linearity range was $10^3$ to $10^6$ copy numbers per reaction for all targets

2 LOD, copy number of the artificial template per gram of feces, equiv. no. of copies (equivalent copy numbers per 1 μL of volume).

3 Coefficients of variance (CVs) at both low and high concentrations are shown.

**Table S2:** Household demographic characteristics by community

| | Total (n=35) | Rural (n=10) | Intermediate (n=15) | Urban (n=10) |
|---|---|---|---|---|
| **Mean maternal age in years (n)** | 26 | 24 (6) | 27 (6) | 26 (5) |
| **Mean child age in months (n)** | 13 | 14 (5) | 12 (6) | 13 (7) |
| **Child sex (%)** | | | | |
| *Male* | 13 (37.1) | 3 (30.0) | 6 (40.0) | 4 (40.0) |
| *Female* | 22 (62.9) | 7 (70.0) | 9 (60.0) | 6 (60.0) |
| **Drinking water source (%)** | | | | |
| *Purchased water* | 16 (45.7) | 0 (0.0) | 11 (73.3) | 5 (50.0) |
| *Well or tubewell water* | 1 (2.9) | 0 (0.0) | 1 (6.7) | 0 (0.0) |
| *Piped water* | 7 (20.0) | 0 (0.0) | 2 (13.3) | 5 (50.0) |
| *Rainwater* | 8 (22.9) | 7 (70.0) | 1 (6.7) | 0 (0.0) |
| *River water* | 3 (8.6) | 3 (30.0) | 0 (0.0) | 0 (0.0) |
| **Bathroom access (%)** | | | | |
| *Household toilet or latrine* | 29 (82.9) | 5 (50.0) | 14 (93.3) | 10 (100.0) |
| *Public or community latrine* | 2 (5.7) | 2 (20.0) | 0 (0.0) | 0 (0.0) |
| *Neighbor's toilet or latrine* | 3 (8.6) | 3 (30.0) | 0 (0.0) | 0 (0.0) |
| *Hole or pit* | 1 (2.9) | 0 (0.0) | 1 (6.7) | 0 (0.0) |

**Table S3:** Enteric pathogen co-occurrence patterns in different animal type.

| Co-ocurrence patterns | Animal type | | | | | | | | |
|---|---|---|---|---|---|---|---|---|---|
| | **Cats** | **Chickens** | **Cows** | **Dogs** | **Ducks** | **Horses** | **Parrots** | **Pigs** | **Total n (%)** |
| aEPEC + *C. parvum.* | 0 | 1 | 0 | 0 | 2 | 6 | 0 | 0 | 9 (16%) |
| aEPEC + *Salmonella spp.* | 0 | 2 | 1 | 1 | 1 | 0 | 0 | 6 | 11 (19%) |
| aEPEC + *C. jenuni/coli* | 0 | 4 | 0 | 3 | 0 | 0 | 0 | 1 | 8 (14%) |
| *Salmonella spp.* + *C. jejuni/coli* | 1 | 2 | 0 | 1 | 0 | 0 | 0 | 0 | 4 (7%) |
| *Salmonella spp.* + *C. parvum* | 0 | 1 | 0 | 0 | 0 | 0 | 0 | 0 | 1 (2%) |
| *Salmonella spp.* + STEC | 1 | 2 | 5 | 2 | 0 | 0 | 0 | 2 | 12 (21%) |
| *C. jejuni/coli* + STEC | 0 | 1 | 0 | 0 | 0 | 0 | 0 | 0 | 1 (2%) |
| *C. parvum* + STEC | 0 | 1 | 1 | 0 | 0 | 2 | 0 | 0 | 4 (7%) |
| *Salmonella spp.* + aEPEC + *C. parvum* | 0 | 0 | 0 | 0 | 1 | 0 | 0 | 1 | 2 (4%) |
| *Salmonella spp.* + *C. jejuni/coli* + aEPEC | 0 | 1 | 0 | 1 | 0 | 0 | 0 | 0 | 2 (4%) |
| *Salmonella spp.* + *C. jejuni/coli* + STEC | 1 | 1 | 0 | 1 | 0 | 0 | 0 | 0 | 3 (5%) |
| **Total n (%)** | 3 (5%) | 16 (28%) | 7 (12%) | 9 (16%) | 4 (7%) | 8 (14%) | 0 (0%) | 10 (16%) | 57 |

**Supplementary Figures**

**Figure S1:** Mean standard curves of gblock 10-fold serial dilutions generated from individual standard curves per gene**.** Concentrations ranged from 10 3 to 10 6 gene copies. error bars represent the standard deviation of Cq values at each concentration.

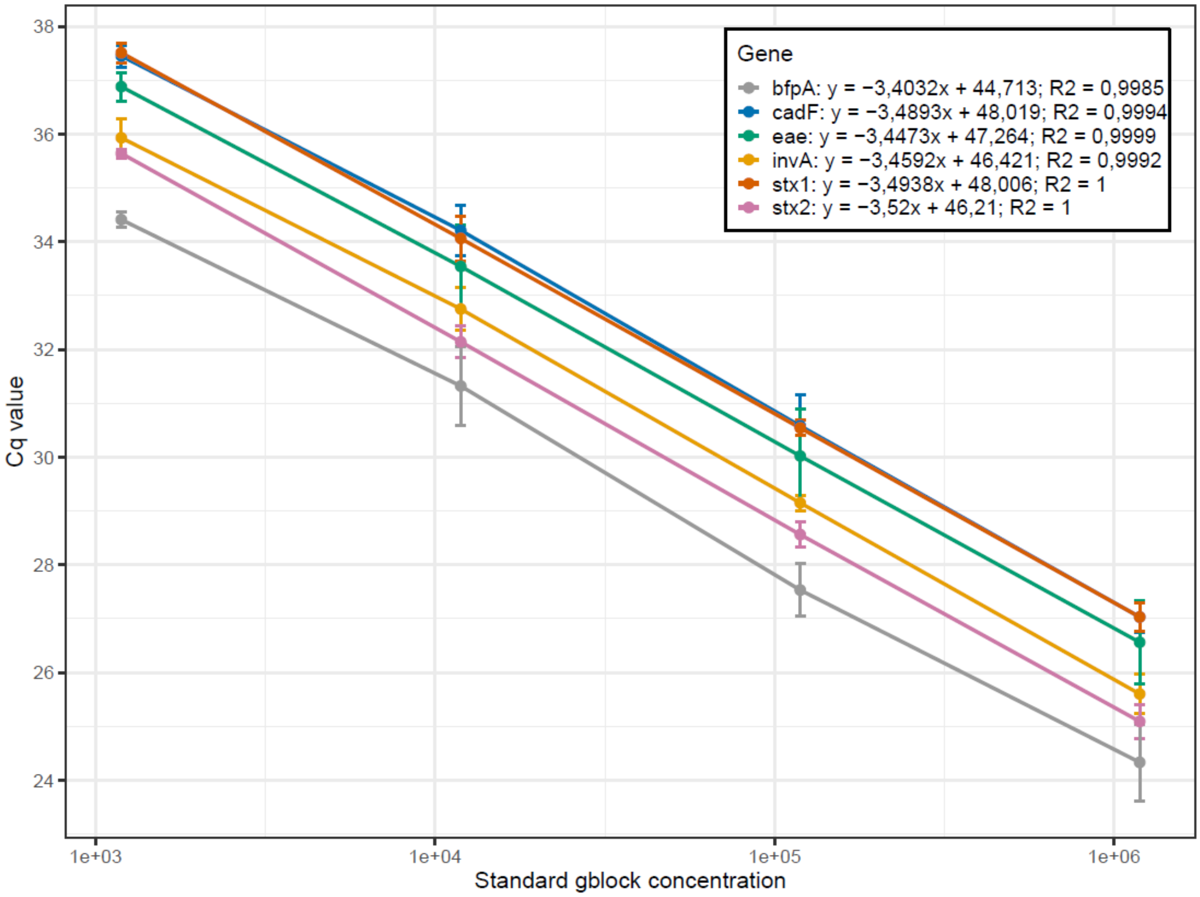

**Figure S2.** Probabilistic co-occurrence model. Heat map showing the random and non-random (negative) enteric pathogen associations determined by the probabilistic co-occurrence model. Enteric pathogens names are positioned to indicate the column and rows that represent their pairwise relationships with other microorganisms

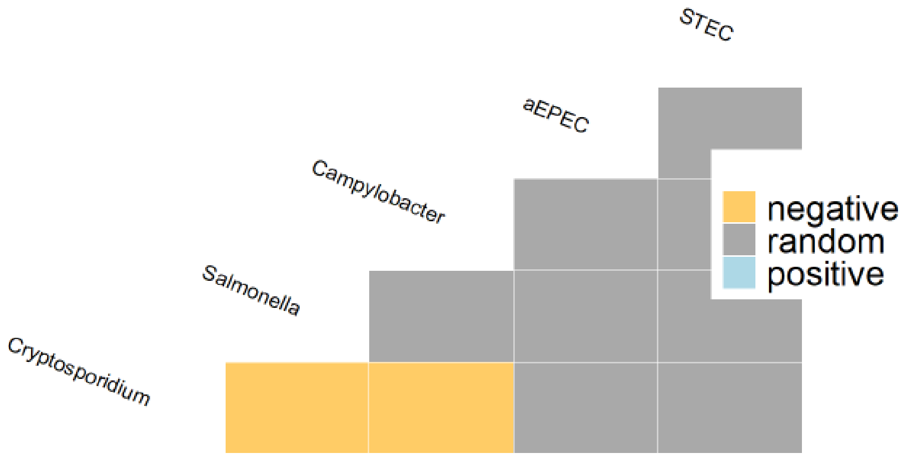
